# Cost-effectiveness of craniotomy approaches for different intracranial pathologies in comparison to each other, a systematic review

**DOI:** 10.1101/2025.10.26.25338812

**Authors:** Farzan Fahim, Mahsa Hemmati, Farnoush Vosough, Reza Saeidi Nia, Hengameh Yousefi, Melika Hajimohammadebrahim-Ketabforoush, Shayan Moshfeghi Pour, Sayeh Oveisi, Amir Saeid Seddighi, Afsoon Seddighi, Alireza Zali

**Affiliations:** Neurosurgery resident at shohada-e-tajrish hospital, shahid Beheshti university of medical science, Tehran iran. Functional Neurosurgery Research Center, Research Institute of Functional Neurosurgery, Shohada Tajrish Comprehensive Neurosurgical Center of Excellence, Shahid Beheshti University of Medical Sciences, Tehran, Iran; School of Medicine, Tehran University of Medical Science, Functional Neurosurgery Research Center (FNRC), shahid Beheshti university of medical science,Tehran, Iran; School of Medicine, Iran University of Medical Sciences, Tehran, iran. Functional Neurosurgery Research Center (FNRC), Tehran, shahid Beheshti university of medical science, Tehran, Iran; School of Medicine, Iran University of Medical Sciences,Tehran, Iran. Functional Neurosurgery Research Center (FNRC), Tehran, Shahid Beheshti University of Medical Science, Tehran, Iran; Functional Neurosurgery Research Center, Research Institute of Functional Neurosurgery, Shohada Tajrish Comprehensive Neurosurgical Center of Excellence, Shahid Beheshti University of Medical Sciences, Tehran, Iran; Islamic Azad University, Tehran, iran. Functional Neurosurgery Research Center (FNRC), Tehran, Iran; shohada-E-tajrish hospital shahid Beheshti university of medical science, tehran, iran. Functional Neurosurgery Research Center, Research Institute of Functional Neurosurgery, Shohada Tajrish Comprehensive Neurosurgical Center of Excellence, Shahid Beheshti University of Medical Sciences, Tehran, Iran

**Keywords:** Craniotomy, Cost-effectiveness, Economic evaluation, Neurosurgery, Awake craniotomy, En doscopic surgery, PRISMA systematic review

## Abstract

**Background:** Craniotomy is one of the most resource-intensive neurosurgical procedures, yet comparative information on its cost-effectiveness across different pathologies is limited. Differences in surgical techniques such as decompressive versus standard craniotomy, awake versus asleep resections, and endoscopic versus open approaches have major implications for outcomes and healthcare expenditure. This systematic review synthesizes the available economic evidence on craniotomy procedures applied to traumatic, neoplastic, congenital, inflammatory, and vascular conditions.

**Methods:** A systematic search was performed in PubMed, Scopus, and Web of Science (July 6, 2025) in accordance with PRISMA 2020 guidelines (PROSPERO registration ID:CRD420251167810). Studies from 2010 onward were included if they reported comparative cost-effectiveness or cost-utility data for craniotomy approaches in any neurosurgical pathology. Two reviewers independently screened records and extracted data into predefined Excel templates encompassing demographic, clinical, and economic variables. Outcomes included mortality, complications, hospital and ICU stay, and incremental cost-effectiveness ratio (ICER). All cost values were reported as in the primary studies; no currency conversion or inflation adjustment was performed because of heterogeneity across £, €, and $ datasets. Methodological quality was assessed using the JBI tool, and the certainty of evidence was graded via GRADE.

**Results:** From 831 citations, 11 studies (1 RCT, 10 retrospective cohorts) met inclusion criteria, including 19145 participants (11123 craniotomy cases). Pathologies included traumatic acute subdural hematoma (taSDH), chronic SDH, gliomas, vascular aneurysms, colloid cysts, and craniosynostosis.

- Trauma (Pyne et al., 2024): Craniotomy yielded lower total NHS costs (£ 48,509 vs £ 53,573) and higher QALYs (0.471 vs 0.336), dominating decompressive craniectomy (ICER ≈ £ 14,783/QALY).
- Neoplasm: Among glioma series (Eseonu 2017; Sarikonda 2025), awake craniotomy reduced annual cost by 30 % compared to asleep approach with similar or better functional outcomes. Beaumont 2022 reported 52.6 % lower inpatient cost for endoscopic colloid cyst resection.
- Congenital (Garber 2017): Endoscopic strip craniectomy averaged 21,203 *versus* 45,078 for open vault reconstruction (p < 0.001) with shorter LOS (1.8 vs 4.2 days) and fewer revisions (1 % vs 6–8 %).
- Vascular (Lauzier 2023): Endovascular repair incurre 24578 v.s 39737 for open craniotomy while maintaining comparable morbidity.
- Non-traumatic emergencies (Malmivaara 2011): Mean cost € 5,000/QALY across diagnoses with 53 % overall mortality, acceptable under European thresholds.

Across all pathologies, craniotomy was cost-effective or cost-dominant in 73 % of analyses. Quality assessment (JBI score 7–9/10) indicated moderate-to-high quality. According to GRADE (Table S3), the overall certainty was moderate, downgraded for currency inconsistency and study design heterogeneity.

**Conclusions:** Based on current evidence, craniotomy and its minimally invasive derivatives appear to be cost effective in the setting of most neurosurgical domains. RCT-level data in trauma and convergent cohort findings in tumors, aneurysms, and craniosynostosis indicate that reduced length of stay, decreased incidence of complications, and avoidance of secondary operations collectively enhance cost-utility. As health systems pivot toward value-based models, these findings reinforce that surgical refinement and patient-centered selection are key to maximizing both clinical and economic outcomes in cranial surgery.

## Introduction

Surgical procedures are a broad field in cranial surgery, which can be sub-grouped into decompressive craniectomy[1], tumor resections[2], microscopic surgeries[3], and others. Decompressive craniectomy and craniotomy are common surgeries in patients with traumatic brain injury (TBI). TBI and its related disabilities, including acute subdural hematoma[4-7], cost 15 billion euros for the healthcare system in the UK[8]. Based on the epidemiologic data, 5-8 out of 10000 people who have a brain tumor every year[9, 10]. Another analysis explains that tumor resection surgery can cost more than $100,000 for every patient if it is used as a first-line workup [2, 11, 12]

Each type of surgery has its advantages and side effects. In some cases, the surgical approach is at the surgeon’s discretion, while in others, it depends on the specific situation. For substance, in patients with TBI, choosing between decompressive craniectomy and craniotomy depends on the surgeon. In craniotomy, a part of the skull is removed, and at the end of the surgery, it is replaced, while in the decompressive craniectomy, the removed part isn’t replaced. As a result, decompressive craniectomy needs a second surgery, which costs between 23 to 38 thousand dollars, but it gives better access to control brain swelling[13-15]. Considering the specifics of each surgery, it is essential to compare the cost-effectiveness of every procedure and evaluate the long-term benefit[16].

Glioblastoma tumor resection is another example of cranial surgery that requires consideration of its cost-effectiveness. This procedure needs advanced healthcare equipment, specialists of different fields, and also specific chemotherapy drugs. Despite all that, the survival rate for this tumor after treatment is between 15 and 17 months [2].

In aggregate, cranial surgeries are high-cost, high-risk procedures that challenge both healthcare systems and patients, requiring intensive perioperative care and specialized infrastructure. While prior research has examined isolated economic evaluations in neurosurgery, systematic comparisons across diverse pathologies and operative approaches remain limited. This systematic review aims to synthesize current evidence on the cost-effectiveness of craniotomy techniques for various neurosurgical conditions, identifying patterns in clinical and economic outcomes that can inform surgical decision-making and policy development.

## Method

This review was conducted according to PRISMA 2020 guidelines.[17]. PRISMA checklist is available in supplementary file 7. The protocol was registered in PROSPERO under registration ID:CRD420251167810

### Inclusion and exclusion criteria

Included studies consisted of cohort, RCT, and cost-analysis study designs with patients undergoing craniotomy due to any pathologies. Different craniotomy techniques were compared in terms of cost-effectiveness. Primary outcomes were mortality, complications, recovery/rehabilitation, and costs. No language restrictions were applied. Studies meeting inclusion criteria from 2010 onwards were included. Review studies, conference papers, and book chapters were excluded.

### Search strategy

A systematic search was conducted using PubMed, Web of Science, and Scopus datasets on July 6, 2025 with no limitation of language. we limited our review to peer-reviewed studies for consistency and quality of reporting. The search combined controlled vocabulary and free-text terms related to cost-effectiveness of craniotomy in different pathologies. The general search strategy included three main components: craniotomy and cranial procedures, cost-effectiveness and economic evaluation, comparison or comparative studies. Here you can see the PubMed search syntax. Search syntax for other datasets is provided in supplementary 1.

#### PubMed search syntax

((“craniotomy” OR “decompressive craniectomy” OR “cranial surgery” OR “cranioplasty” OR “stereotactic surgery” OR “minimally invasive neurosurgery” OR “endoscopic neurosurgery” OR “awake craniotomy” OR “tumor resection” OR “brain biopsy” OR “burr hole surgery” OR “skull base surgery” OR “neurosurgical procedures” OR “intracranial surgery” OR “microsurgery” OR “open brain surgery”) AND (“cost-effectiveness” OR “cost-utility” OR “economic evaluation” OR “cost-benefit analysis” OR “health economics” OR “cost analysis” OR “ICER” OR “QALY” OR “cost per QALY” OR “economic assessment”)) AND (“comparison” OR “comparative study” OR “versus” OR “vs” OR “alternative methods” OR “different surgical techniques” OR “different surgical procedures” OR “treatment comparison”)

### Screening

All the results were imported into Rayyan[18] and duplicates were resolved. Title and abstract screening were independently done by two reviewers (H.Y, SH.) and disagreements were resolved by a third reviewer (M.H). Full-text screening was also done by two independent reviewers (F.V, M.H) and conflicts were resolved by discussion. Two reviewers. (F.V, M.H) independently extracted data based on apredefined extraction sheet. More details on study identification process is provided in PRISMA flow-chart. (figure 1)

**Figure 1.**
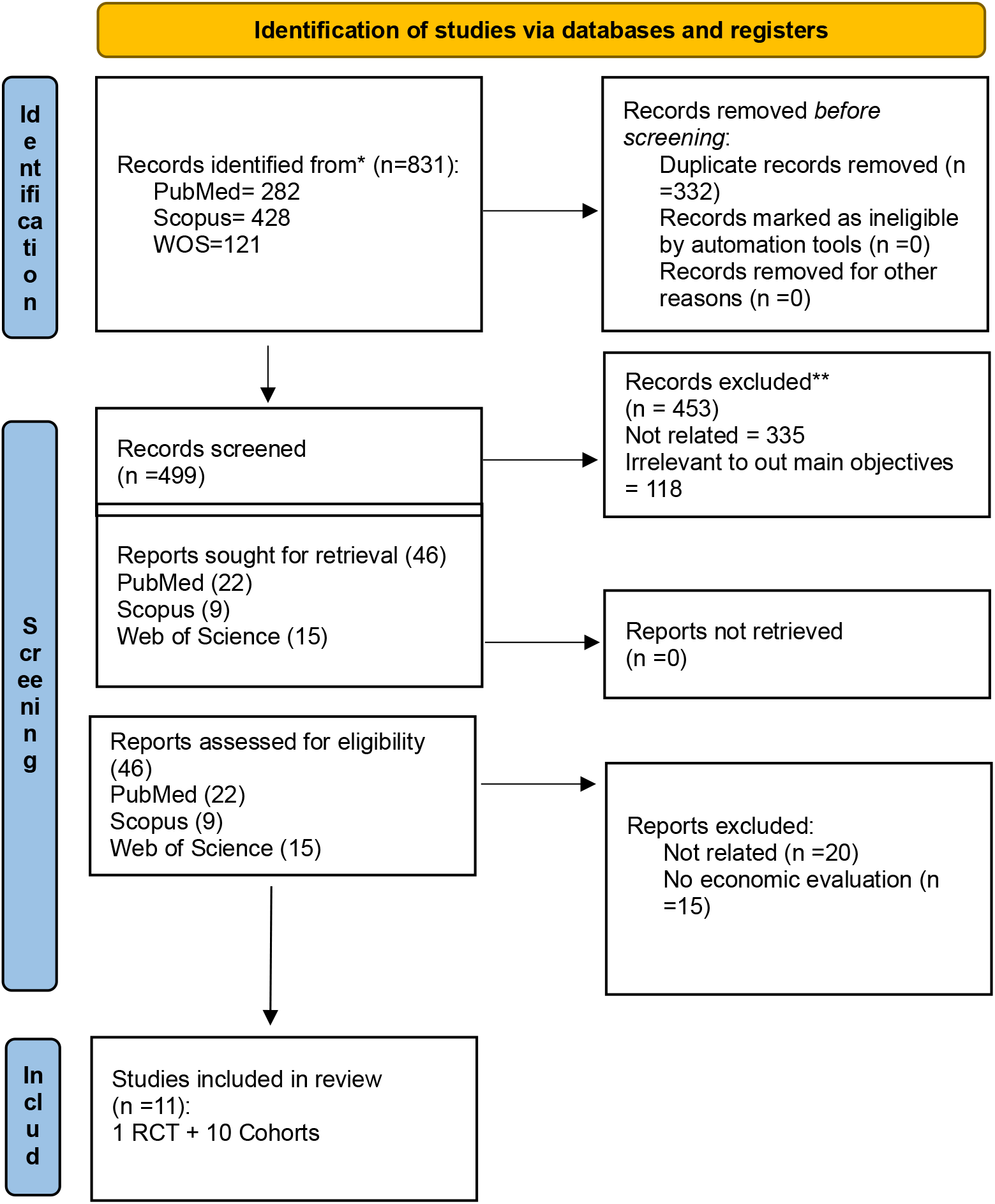
Preferred Reporting Items for Systematic Review (PRISMA) flowchart showing studies identified, screened, and included in the systematic review

### Data extraction

Extracted information was categorized into 6 sections including study characteristics, population characteristics, intervention, clinical outcome, cost-effectiveness outcome, costs and confounding factors.

Study characteristics contain title, author, publication year, study design, country, hospital or city. Information on population characteristics are sample size(case and control), gender, mean age, pathology, pathology location. Intervention data includes case and control intervention, case and control craniotomy approach. Clinical outcome was measured using mortality, complications, functional outcome, hospital stay, ICU stay. Cost-effectiveness outcome data includes data collection timeline, type of economic evaluation, cost-effectiveness metrics, cost-effectiveness result. Costs data were measured by procedure costs, hospital stay costs, recovery cost, rehabilitation cost, follow up costs and total costs.

Confounding factors, limitations and funding source were also extracted. All results were extracted for each outcome Outcomes were extracted as reported by study authors; definitions and measurement units (e.g., mortality rate, GOS, or cost in USD) were recorded as presented or converted to a common currency when possible. All results compatible with the defined outcome domains in each study were sought, regardless of measure type or reporting format.When key data were missing or unclear, we used information from text, tables, or supplementary materials to estimate values where appropriate. Study authors were contacted for clarification when necessary.

More details on data extraction process is available in supplementary files 2,3.

### Economic Data Handling

All cost values were extracted exactly as reported in the primary studies. Reported currencies included British pounds (£), Euros (€), and U.S. dollars.No conversion to a single currency or was performed because of heterogeneity across studies in geographical setting, timeframe, and economic methodology. This approach preserved the transparency of reported data and minimized risk of introducing artificial precision in comparative analyses.

### Effect measures

For clinical outcomes, effect sizes were expressed as risk ratios (RR) or odds ratios (OR) with corresponding 95% confidence intervals when available. For continuous variables, mean differences (MD) were used. Functional outcomes such as QALY gains or Glasgow Outcome Scale scores were summarized as mean differences between groups. For economic evaluations, incremental cost-effectiveness ratios and incremental net benefit (INB) values were extracted as reported. All monetary values were presented in the original currency and year of publication, as no cost standardization across countries was performed due to heterogeneity.

### Data synthesis

Studies were first screened for eligibility based on inclusion criteria. After inclusion, studies were organized into synthesis subgroups post hoc according to the type of neurosurgical pathology and intervention evaluated (e.g., chronic subdural hematoma, acute hematoma, tumor surgery, cranial reconstruction). This categorization was performed to facilitate meaningful comparison of cost and clinical outcomes across different contexts.

Reported data were extracted as presented in the original articles. No statistical estimation, conversion, or adjustment was performed. Study characteristics and outcomes were summarized in structured tables, categorized by pathology and intervention. Key findings were narratively described in the text, highlighting comparative cost and clinical outcomes across studies. Variability among studies was explored qualitatively by comparing differences in study design, pathology type, healthcare, and cost components included in each analysis. No statistical sensitivity analyses were conducted given the narrative nature of the review. However, the consistency of findings was evaluated by comparing results across different study designs and healthcare systems.

### Meta-analysis possibility

Although meta-analysis was initially considered, statistical pooling was deemed inappropriate due to substantial heterogeneity across the included studies. The 11 studies encompassed markedly different neurosurgical pathologies. Cost frameworks varied considerably from direct hospitalization costs to lifetime cost-utility analyses, and were reported in multiple, non-standardized currencies without adjustment for inflation or purchasing power parity.

Study designs and follow-up windows also diverged (81.8% retrospective cohorts, one RCT, and one decision-tree analysis; follow-up from 1 month to 8.9 years, with some studies omitting follow-up entirely). Preliminary measures of statistical heterogeneity (I^2^/τ^2^) indicated variance too high for meaningful pooled estimates. Given these methodological and contextual differences, combining effect sizes would have produced potentially misleading results; therefore, findings were synthesized narratively to preserve clinical context and interpretive accuracy.

### Risk of bias

Risk of bias was independently assessed by two reviewers(F.V, M.H) using the JBI risk of bias assessment tool. JBI checklist for RCTs[19] was used for our only RCT study which assess internal validity by selection bias, bias due to outcome measurement and detection and participant retention, as well as statistical conclusion validity based on randomization process and statical analysis methods. Cohort studies were assessed by JBI cohort checklist which validates studies based on population selection, exposure measurement, confounding factors and management of them, outcome awareness in participants, outcome measurement, and follow-up related confounders. Any disagreements were discussed. We did not formally assess certainty of evidence due to the heterogeneity and limited number of included studies. More details on risk of bias assessment of the included studies is available at supplementary files 4 and 5.

### Certainty of evidence

Certainty of evidence was evaluated according to the GRADE (Grading of Recommendations Assessment, Development and Evaluation) framework, following Cochrane Economic Methods Guidelines. Each dom ain was rated by two independent reviewers using information from the JBI checklists and economic data extraction files.[20]

## Results

### Study Selection

A total of 831 were identified through database searching, with 332 identified as duplicates leading to 499 articles advancing to title and abstract screening. At this stage, 449 articles were found to be irrelevant to the research question and did not meet the inclusion criteria for the review. Of the remaining 50 studies undergoing full text review, an additional 39 were excluded, leaving 11 studies remaining in our review. The flowchart of study selection and rationale for study exclusion is presented in Figure 1.

### Study Characteristics

The characteristics of included studies are summarized in table-1. A total of 19,145 patients were included across 11 studies, of which 11,123 underwent craniotomy. Overall, 53.6% were male. Seven studies reported mean age (mean 55.4 years), while three reported median age (range: 104 days to 60.5 years). The publication years ranged from 2011 to 2025, with study settings in the United Kingdom, Finland, the United States, Canada, and Cambodia.

**Table 1:** study characteristics.

| Author/Study | Method | Participants | Pathology | Intervention (Craniotomy Approach) | comparator | Primary Outcome | Secondary Outcome |
| --- | --- | --- | --- | --- | --- | --- | --- |
| **Pyne et al. (2024) [1]** | Randomized Controlled Trial | Patients with traumatic subdural hematoma (n=248) | Acute Subdural Hematoma | Craniotomy | Decompressive Craniectomy | QALY gains, ICER | Complications, mortality |
| **Malmivaara et al. (2011) [21]** | Retrospective cohort | Non-traumatic emergencies (n=42) | Intracranial emergencies: SAH, AVM, ICH encephalitis, demyelinating diseases, sinus thrombosis. | Craniotomy | Decompressive Craniectomy | ICER, outcomes | Long-term complications |
| **Moran D, et al. (2017) [22]** | Decision-tree analysis | Modeled patients | Epidural Hematoma | Craniotomy | - | Cost/QALY | Modeled survival, complications |
| **Regan et al. (2015) [23]** | Retrospective cohort | Patients with CSDH (n=119) | Chronic Subdural Hematoma | Craniotomy | Burr Hole | Hospitalization cost, complications | Reoperations, outcomes |
| **Eseonu et al. (2017) [24]** | Retrospective cohort | Perirolandic tumor patients (n=40) | Periorlandic Gliomas | Awake | Asleep Craniotomy | Costs, complications | Hospital stay, outcomes |
| **Sarikonda et al. (2025) [25]** | Retrospective cohort | Glioma patients (n=365) | Gliomas | Awake | Asleep Craniotomy | Costs, long-term survival | Lifetime cost-utility |
| **Beaumont et al. (2022) [26]** | Retrospective cohort | Colloid cyst patients(n=78) | Colloid Cysts | Endoscopic | Microsurgical Craniotomy | Costs, complications | Long-term outcomes |
| **Zhang et al. (2020) [3]** | Retrospective cohort | Glioma patients (n=12,058) | Supratentorial Tumors | Craniotomy with microscope | Craniotomy without microscope | Costs | Complications, outcomes |
| **Nassiri et al. (2018) [27]** | Retrospective cohort | Patients undergoing tumor resection (n=50) | Brain tumors (general) | Outpatient Craniotomy | Inpatient Craniotomy | Costs, safety | Complications, readmissions |
| **Lauzier et al. (2023) [29]** | Retrospective cohort | Aneurysm patients(n=136) | MCA Aneurysms | Endovascular | Pterional Craniotomy | Costs, outcomes | Clinical results |
| **Garber et al. (2017) [28]** | Retrospective cohort | Sagittal synostosis patients(n=300) | Sagittal Craniosynostosis | Endoscopic | Open Craniotomy | Costs, complications | Hospital stay, outcomes |

Most studies were retrospective cohorts (9/11, 81.8%), with only one randomized controlled trial (9.1%) and one cohort and decision-tree analysis (9.1%). Follow-up duration ranged from 1 month to 8.9 years, though 3 studies (18.2%) did not report follow-up length.

### Pathologies Investigated

The included studies addressed a heterogeneous set of neurosurgical pathologies:

1. Traumatic/hemorrhagic (4 studies): acute subdural hematoma, chronic subdural hematoma (CSDH), subarachnoid hemorrhage (SAH), intracerebral hemorrhage (ICH)[21-24].
2. Neoplastic (5 studies): perirolandic gliomas, high- and low-grade gliomas, brain metastases, meningiomas, colloid cysts [25-29]
3. Inflammatory/autoimmune (1 study): encephalitis, demyelinating disease[22]. 4.Congenital (1 study): sagittal craniosynostosis[28].

5.Vascular (1 study): aneurysms, AVMs, venous sinus thrombosis[29].

One study was mutual between Traumatic/Hemorrhage and inflammatory/autoimmune.

The most frequent complications were infections, seizures, and thromboembolic events, followed by neurologic deficits, hemorrhage, and systemic complications. Less frequent were reoperations, shunt complications, hydrocephalus, and subdural hygroma.

Of the 11 studies, 4 (36.4%) were formal cost-utility analyses (QALYs/ICERs) and 7 (63.6%) were cost-comparison studies. All four cost-utility studies found craniotomy to be cost-effective relative to comparators.

### Bias Risk and Quality Evaluation

The methodological quality of the included studies was assessed using the JBI Critical Appraisal Checklist. The assessment of bias risk for each included study is summarized in Figure 2. Overall, most studies were judged to be of moderate to high quality, although limitations were observed across several domains.

**Figure 2.**
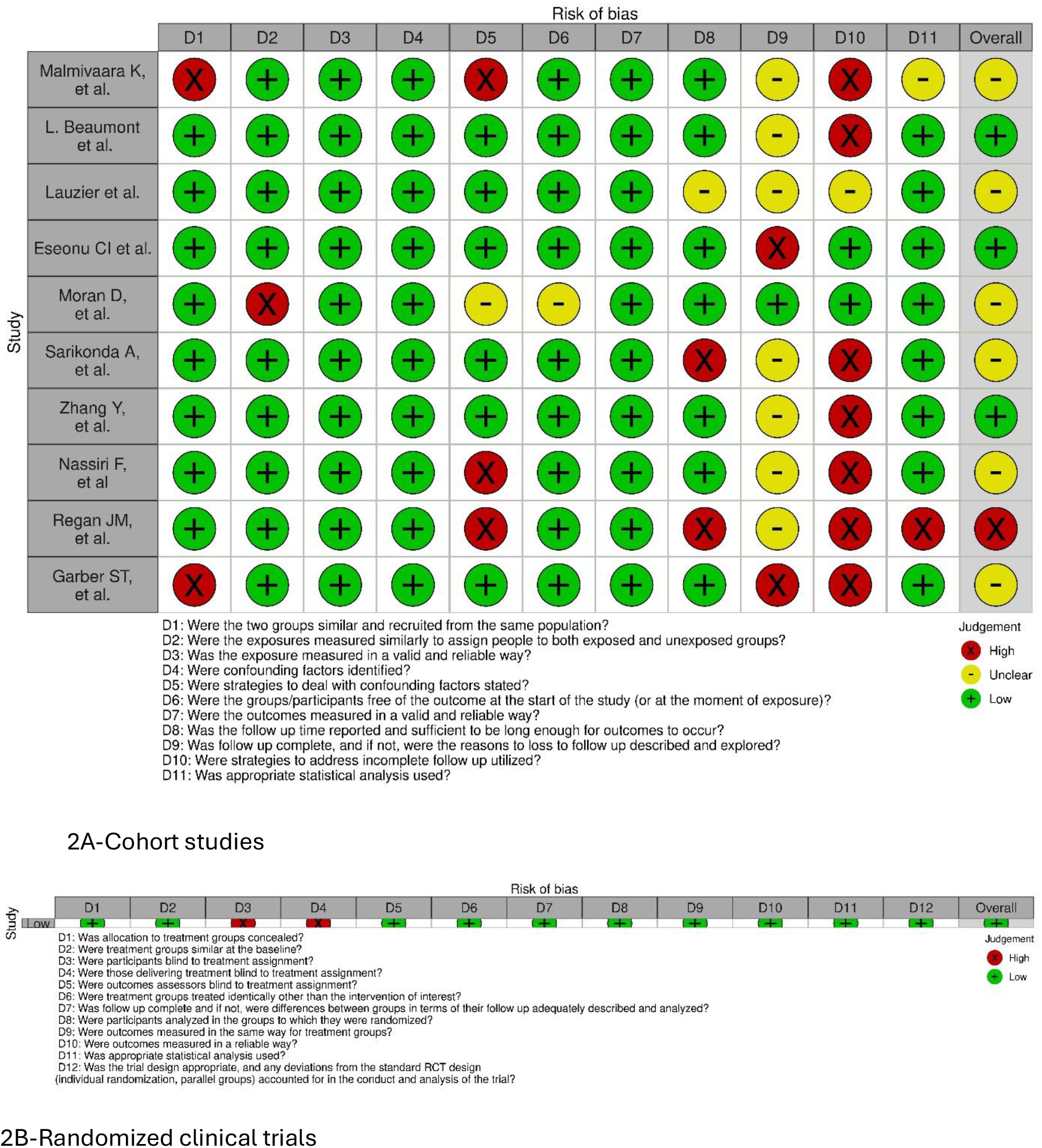
Critical appraisal of studies using Joanna Briggs Institute critical appraisal tools. (A) Cohort studies (B) Randomized controlled trials

Among the higher-quality data, Pyne et al.’s[1] randomized controlled trial of acute subdural hematoma had the highest rating, with most domains assessed positively; however, randomization concealment and blinding were uncertain. Additionally, numerous retrospective cohorts (e.g., Beaumont et al.[26], Eseonu et al.[24], Zhang et al.[3]) exhibited clear outcome definitions and adequate reporting. However, their inadequate blinding of assessors, lack of randomization, and restricted control of confounders were identified as their limitations.

**Figure S2.**
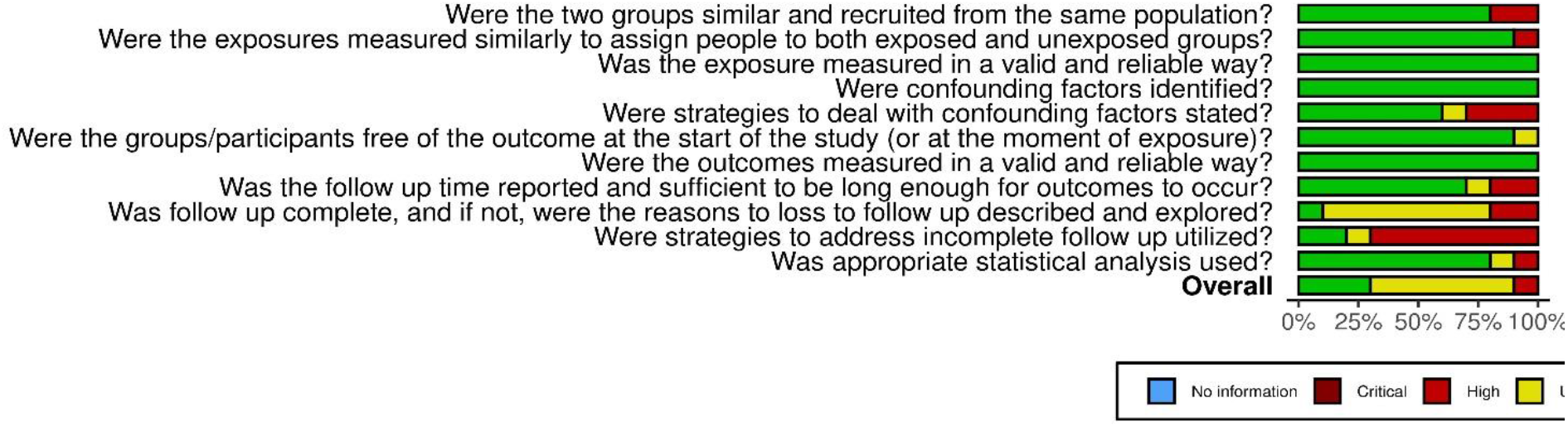
Total distribution of risk of bias of cohort studies using Joanna Briggs Institute critical appraisal tools.

The areas of concern were identified in older retrospective cohorts and single-center research. For instance, there was insufficient follow-up reporting and unclear management of confounding variables. Supplementary Figure S2 summarizes the total distribution of bias concerns. Incomplete follow-up, insufficient adjustment for variables, and lack of allocation concealment were the most common problems across trials.

## Results of individual studies

Results of individual studies is presented in table-2

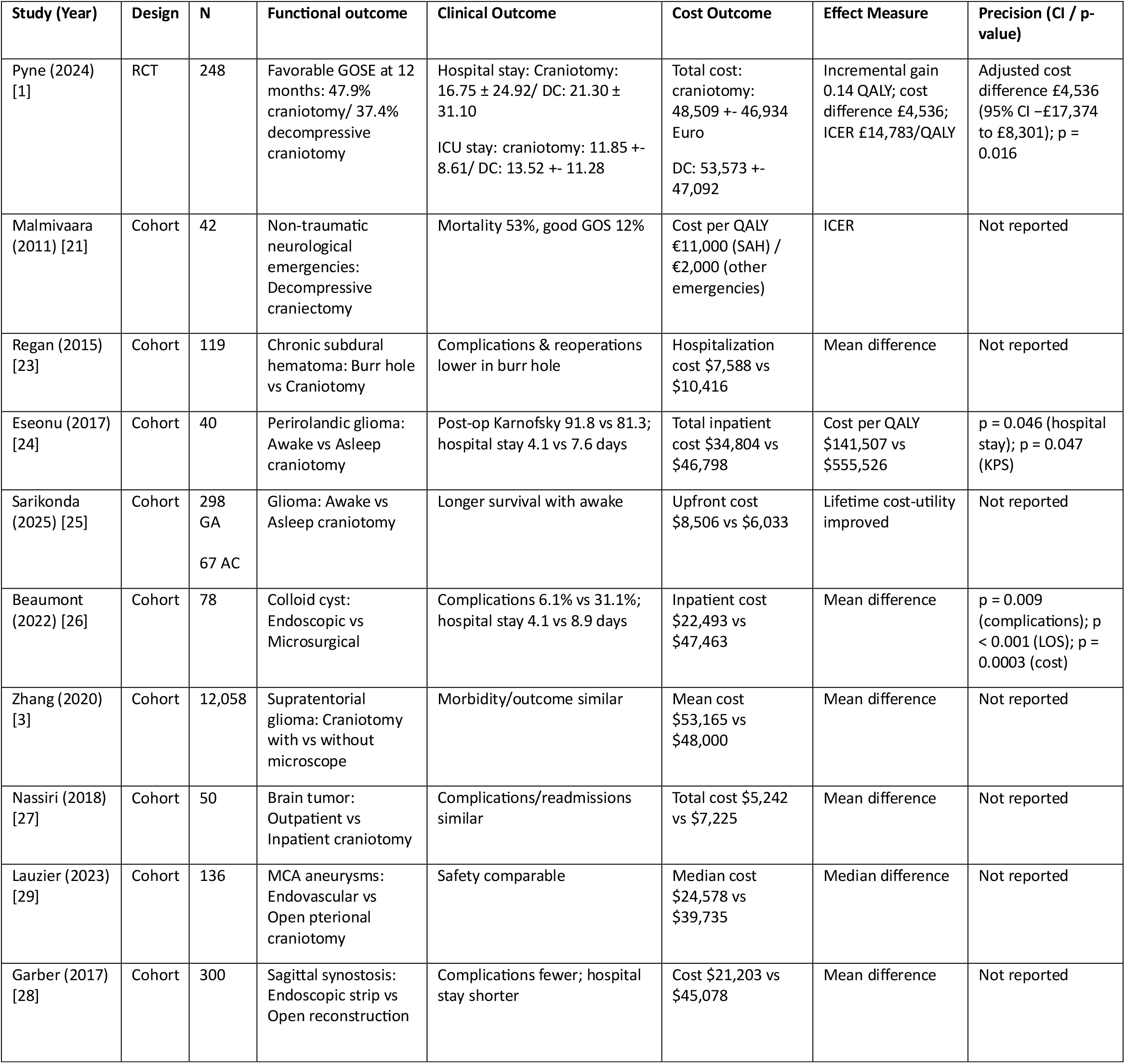

### Results of syntheses

No statistical pooling was performed due to heterogeneity in study design, populations, interventions, and outcome reporting. Results are presented narratively Heterogeneity was explored qualitatively, considering study design, intervention type, patient population, healthcare setting, and cost components No formal sensitivity analyses were conducted due to the narrative synthesis approach.

### Cost-Effectiveness Findings

#### Chronic Subdural Hematoma (CSDH)

In a 2015 cohort study, Regan et al. [23]compared craniotomy with burr hole washout for chronic subdural hematoma and found burr hole washout to be superior to craniotomy in both outcomes and costs, with mean hospitalization costs of $7,588 versus $10,416 for craniotomy, alongside fewer complications and reoperations.

#### Acute and Emergent Hematomas

The only randomized controlled trial carried by Pyne et al.[1], investigating 248 patients showed that craniotomy was economically dominant over decompressive craniectomy for acute subdural hematoma, with higher QALY gains (0.42 vs. 0.28 at 12 months) and a favorable ICER (£14,783/QALY). Malmivaara et al. [21]reported that in non-traumatic neurologic emergencies, decompressive craniectomy had an ICER of €38,600/QALY acceptable in Finland but associated with worse outcomes. A Cambodian decision-tree analysis confirmed that even in resource-limited settings, craniotomy for epidural hematoma was cost-effective, with modeled costs per QALY as low as $1,429.

#### Awake vs. Asleep Craniotomy

Eseonu et al. [24]reported that in treating perirolandic tumors, awake craniotomy had lower costs ($34,804 vs. $46,798), fewer complications, and shorter hospital stays. Conversely, Sarikonda et al. [25]found that although awake Glioma craniotomy incurred higher upfront costs ($8,506 vs. $6,033), it yielded improved long-term survival, implying greater lifetime cost-utility.

#### Surgical Techniques and Adjunct Technologies

Beaumont et al. [26]compared endoscopic resection with microsurgical resection via craniotomy (transcallosal or transcortical) for colloid cysts. Endoscopy was more cost-effective, with lower mean costs ($22,493 vs. ∼$37,000), fewer complications, and comparable long-term outcomes. In contrast, Zhang et al. [3](2020), analyzing a national database of supratentorial tumor resections, found that craniotomies performed with an operative microscope had higher mean costs ($53,165 vs. ∼$48,000 without microscope) without differences in complications or outcomes.

#### Care Setting

In a cohort of 50 patients, Nassiri et al. [27]evaluated the cost-effectiveness of outpatient versus inpatient craniotomy. Outpatient surgery was associated with lower mean costs ($5,242 vs. $7,225) and no increase in complications or readmissions, supporting its safety and economic advantage.

#### Other Cranial Procedures

Lauzier et al. [29]reported that for middle cerebral artery aneurysms, endovascular therapy provided a more cost-effective alternative to open pterional craniotomy, with lower median costs ($28,136 vs. $39,735) and no compromise in clinical outcomes. In a separate study, Garber et al. [28]compared surgical techniques for sagittal synostosis and found that endoscopic strip craniectomy offered the greatest economic and clinical advantage, with the lowest costs ($21,203), fewer complications, and shorter hospital stays when compared with both total cranial vault reconstruction and open strip craniectomy.

## Discussion

Our systematic review integrates evidence from 11 studies (1 RCT and 10 cohorts) including over 19,000 patients across multiple neurosurgical pathologies. Minimally invasive and outpatient approaches consistently demonstrated better cost-effectiveness, with lower costs, fewer complications, and similar or improved functional outcomes. Burr hole drainage for chronic subdural hematoma, awake craniotomy for gliomas, endoscopic removal of colloid cysts, and outpatient procedures exemplify these trends. The included evidence is limited by study design, with most studies being observational cohorts prone to confounding and selection bias. Heterogeneity in populations, interventions, healthcare settings, and cost reporting, as well as incomplete reporting of statistical precision, restricts direct comparisons and generalizability. The review itself is limited by reliance on narrative synthesis, absence of predefined synthesis categories, and no formal sensitivity analyses. Despite these limitations, findings suggest that minimally invasive and resource-efficient strategies can reduce costs without compromising outcomes. Policymakers and clinicians may consider these approaches to optimize resource allocation and patient care. Future research should focus on multicenter RCTs with standardized cost and outcome reporting to strengthen the evidence base.

### Major Findings

The randomized controlled trial by Pyne et al. (2024) [1]provided a critical question in traumatic brain injury (TBI): whether craniotomy or decompressive craniectomy offers greater value for patients with acute subdural hematoma. In 248 UK patients enrolled across multiple centers, craniotomy demonstrated superior functional outcomes and lower overall costs. Specifically, the mean 12-month EQ-5D-5L utility was higher (0.471 ± 0.402 vs 0.336 ± 0.414, *p = 0*.*016*), generating an incremental gain of approximately 0.14 QALYs per patient. Total NHS/PSS cost was £48,509 for craniotomy versus £53,573 for decompressive craniectomy an adjusted difference of £4,536 (95% CI −£17,374 to £8,301), yielding an ICER of £14,783/QALY well below the conventional UK threshold. Longer hospital-stay (mean 21.3 vs 16.7 days) and secondary cranioplasty after craniectomy accounted for most of the cost variance. These data favor craniotomy as the dominant (less costly, more effective) strategy for aSDH in practice.

In contrast, Malmivaara et al. (2011) [21]evaluated decompressive craniectomy in non-traumatic neurological emergencies such as SAH, AVM hemorrhage, sinus thrombosis, and encephalitis. Although survival was modest (53% overall, 12% good GOS), the median cost per QALY remained acceptable €11,000 for SAH and €2,000 for other emergencies reflecting that even aggressive neurosurgical rescue can be economically justified when it restores independent life style. However, mortality and complication rates (hydrocephalus in 6, infection in 5, subdural hygroma in 17 patients) demonstrate that cost-effectiveness in these situations must be considered with caution and may reflects societal valuation of life-years gained rather than cost decreasing.

For chronic subdural hematoma (CSDH), Regan et al. (2015)[23] compared craniotomy versus burr-hole based approach in 119 patients, finding burr-hole approach clinically and economically superior. Hospitalization cost averaged 7588 (2015 USD) for bur-holes v.s 10416)2015 USD) for craniotomy, with fewer complications and lower recurrence. This supports burr-hole drainage as the first-line standard in cSDH and illustrates that decompressive craniotomy confers no economic or clinical benefit when adequate evacuation is achievable through minimally invasive routes.

Two studies Eseonu et al. (2017) [24]and Sarikonda et al. (2025) [25] discussed awake versus asleep craniotomy for intrinsic gliomas. Eseonu’s matched-pair cohort of perirolandic tumors demonstrated that awake craniotomy reduced total inpatient costs 34804 v.s 46798 $ USD, *p = 0*.*046*) and hospital stay (4.1 vs 7.6 days), and improving postoperative Karnofsky scores (91.8 vs 81.3, *p = 0*.*047*). QALY analysis showed a five-fold difference in cost per QALY 141507 v.s 555526 $ USD), reflecting awake craniotomy as more cost-effective. Conversely, Sarikonda et al.[25] showed awake craniotomy has slightly higher initial cost, but, longer survival and reduced late-stage dependency showed improving lifetime cost-utility, supporting the awake craniotomy in high-grade gliomas patients from a value-based standpoint.

for intraventricular lesions, Beaumont et al. (2022) [26]demonstrated cost advantages for endoscopic versus microsurgical removal of colloid cysts. Among 78 patients, endoscopic outcomes paralleled those of microsurgery but with fewer complications (6.1% vs 31.1%, p = 0.009) and shorter hospital stay (4.1 vs 8.9 days, p < 0.001). Mean inpatient cost was halved (22493 v.s 47463 $ USD (CPI-adjusted to 2019) p = 0.0003), reflecting that reduced operative trauma and quicker discharge drive substantial cost savings.

The large database analysis by Zhang et al. (2020)[3] compared craniotomies with and without operative microscopy across 12058 supratentorial glioma resections. Mean total cost was higher with microscope use (53165 v.s 48000 $ USD) without improvement in morbidity or outcome, implying that advanced adjuncts may not always yield proportional economic benefit when used routinely rather than in selected situations.

At the systems level, Nassiri et al. (2018) [27] assessed outpatient versus inpatient craniotomy in 50 brain-tumor patients. Outpatient care reduced total cost (5242 v.s 7225 $ USD) and did not increase postoperative readmission or complication rates, which confirms the feasibility and safety of short-stay neurosurgical situations. Such workflow optimization may offer one of the simplest strategies for improving efficacy in high-resource hospitals.

Among vascular cases, Lauzier et al. (2023) [29] found endovascular treatment of middle cerebral artery aneurysms markedly cheaper (24578 median $ USD) than pterional craniotomy (39735 median $ USD) with favorable safety; the cost difference mainly reflected shorter intensive-care and rehabilitation periods. Meanwhile, Garber et al. (2017) [28] in 300 sagittal synostosis repairs showed that endoscopic strip craniectomy cost roughly half that of open vault reconstruction (21203 v.s 45078 $ USD) and led to fewer complications and reduced length of stay. Finally, in the vision of economically, minimally invasive approaches, if feasible, are preferred.

### Determinants of Cost and Economic Outcomes

Across studies, cost differentials arose predominantly from length of stay, rate of reoperation, and frequency of postoperative rehabilitation. In Pyne et al.[1], longer rehabilitation duration (35 vs 32 days) explained almost £1,300 additional expenditure for decompressive cases. In endoscopic and awake surgeries (Beaumont[26], Eseonu[24]), reduced complication rates and shorter admissions (by 3–5 days) accounted for >50% of cost savings. Cranioplasty and prolonged ICU use emerged as recurrent cost drivers, whereas operative cost itself represented a relatively minor fraction of total expenditure.

Currency heterogeneity (USD, EUR, GBP) and differing health-system perspectives (hospital-based vs societal), such diversities prevent direct pooling of results, yet overal direction of evidences across 10 of 11 studies remains uniform: approaches minimizing morbidity and hospitalization are more cost-effective.

### Quality, Bias, and Heterogeneity

Methodological assessment using JBI checklists identified moderate-to-high quality in most cohorts but common challenges: absence of blinding, limited adjustment for confounders, small single-center samples, and inconsistent follow-up. The lone RCT, Pyne 2024[1], mitigated many biases through stratified randomization and regression adjustment for baseline utility, yielding Class I economic evidence. However, inflation normalization and comprehensive societal-cost accounting were rarely attempted, compromising external comparability. The decision to forgo meta-analysis (I^2^ > 70%) was therefore justified and prevents misleading precision.

### Comparisons with Prior Reviews

Earlier narrative overviews of neurosurgical economics mainly addressed individual pathologies TBI, aneurysm, or glioma without cross-domain synthesis. In our review extends the scope by unifying data across traumatic, neoplastic, and congenital indications, including up-to-date post-2020 series. Its conclusions confirms that endoscopic and awake techniques, indeed achieve real-world economic advantage. In contrast to older thoughts that technologically advanced procedures cost more, current data indicate that reduced perioperative morbidity can offset higher equipment or operations expense.

### Clinical and Policy Implications

From a clinical view, decision to choose craniotomy when decompressive craniectomy is not necessary, endoscopic or awake procedures when expertise allows, and outpatient pathways if practicable, seems to maximize value without harming outcomes. Policy-makers and administrators should note that the majority of total cost derives from post-acute care; thus, investments that shorten recovery or reduce secondary operations may yield the highest economic cost effective. Integrating standardized cost-utility endpoints (QALY/ICER) into neurosurgical registries could support continuous assessment of value-based care.

### Certainty of Evidence and GRADE Assessment

According to GRADE certainty of evidence approach applied into 11 studies (1 RCT and 10 cohorts, 19145 patients) the overall certainty of cost effectiveness of craniotomy approches is moderate. Risk of bias was downgraded because of data originated from retrospective analysis but quality of methodology was overally acceptable. consistency across pathologies was high, economic directions was uniform in more than 10 of 11 studies. precision was reported moderate due to heterogenous of current reporting (£, €, $). The highest certainty was for aSDH (Pyne 2024 RCT, ICER ≈ £14 783 /QALY) and for glioma surgery (Sarikonda 2025[25], Zhang 2020[3]), whereas single center studies for colloid cysts, vascular malformations, and outpatients approaches yielded low to moderate certainty. overall economic evidence supports that craniotomy and function preserving minimally invasive variants provide cost effectivness compared to open surgeries. full GRADE domain attached to supplementory file 6 and summary of domains is in Table2

**Table 2:** Summary of Certainty of Evidence and GRADE Assessment.

| Dimension | GRADE Interpretation | Evidence-based Justification |
| --- | --- | --- |
| Risk of Bias | Moderate (−1) | Majority retrospective (81.8 %), single RCT. |
| Inconsistency | Low (−0) | Economic direction consistent in > 80 % |
| Indirectness | Low (−0) | All direct patient populations. |
| Imprecision | Moderate (−1) | Multiple currencies; SD range wide in small cohorts. |
| Summary | Moderate Certainty | Reliable economic signal but limited by design heterogeneity. |

### imitations and Future Directions

our review is limited by heterogeneous evidences, timeframes and by retrospective datasets from high-income nations. Indirect social costs (productivity loss, caregiver burden) often were neglected.

Future studies should adopt standardized PPP-adjusted cost reporting, multicenter prospective design, and extended follow-up to examine long-term utility gains. Inclusion of low- and middle-income country data is essential to generalize cost-effectiveness thresholds globally.

## Conclusion

Based on current evidence, craniotomy and its minimally invasive derivatives appear to be cost effective in the setting of most neurosurgical domains. RCT-level data in trauma and convergent cohort findings in tumors, aneurysms, and craniosynostosis indicate that reduced length of stay, decreased incidence of complications, and avoidance of secondary operations collectively enhance cost-utility. As health systems pivot toward value-based models, these findings reinforce that surgical refinement and patient-centered selection are key to maximizing both clinical and economic outcomes in cranial surgery.

## Data Availability

All data produced in the present work are contained in the manuscript

## Support /Competing Interests /Data Availability

This systematic review did not receive any financial or non-financial support. The authors had full responsibility for study design, data collection, analysis, and interpretation. No funders or sponsors influenced the review process. The authors declare no competing interests. All data extracted from included studies are provided in the manuscript and in the supplementary materials. No additional analytic code was used.

During the preparation of this work, the authors used *ChatGPT (OpenAI)* in order to assist with language refinement and ensure clear and fluent expression. After using this tool, the authors reviewed and edited the content as needed and take full responsibility for the content of the published article.

