## supplementary file 6 for "Cost-effectiveness of craniotomy approaches for different intracranial pathologies in comparison to each other, a systematic review"

**GRADE CERTAINTY of EVIDENCE:**

| **Domain / Pathology** | **Source Studies (n, design, country)** | **Directness (Population & Intervention relevance)** | **Risk of Bias (JBI mean score / 10)** | **Consistency of Findings** | **Precision (95 % CIs / SD)** | **Effect Direction (Economic → Clinical coherency)** | **Certainty (GRADE level A-D)** | **Key Explanation and Data Support** |
|---|---|---|---|---|---|---|---|---|
| Traumatic Acute Subdural Hematoma (ASDH) | Pyne 2024 (RCT, UK HTA) + Malmivaara 2011 (Finland cohort) + Moran 2017 (model, Cambodia) | **High:** similar setting, identical surgery comparison (craniotomy vs DC) | 9.0 (RCT), 7.5 (cohort) | High – all favor craniotomy economically | ICER 95 % CI – £17 374 to £8 301 | Craniotomy dominant (lower mean cost £4 536; higher QALY 0.42 vs 0.28) | **A: Moderate to High** | Only RCT available; good adjustment; moderate imprecision. |
| Non-Traumatic Neurological Emergencies | Malmivaara 2011 (Finland) | **Moderate:** varied etiologies (SAH, AVM, ICH) | 7 / 10 | N/A (single study) | Not reported | ICER ≈ €5 000 / QALY (acceptable threshold Finland) | **B: Moderate** | Heterogeneous sample downgrades consistency but adequate reporting. |
| Chronic Subdural Hematoma (CSDH) | Regan 2015 (USA retrospective n = 119) | High | 7 / 10 | High (burr-hole dominance clear) | Narrow (7 588 $vs$ 10 416) | Burr-hole cheaper and clinically better → high internal alignment. | **B: Moderate** | Only one center but large effect size justifies moderate certainty. |
| Glioma Awake vs Asleep | Eseonu 2017 (USA), Sarikonda 2025 (USA/Canada), Zhang 2020 (China) | High: all address functional preservation glioma | 7.5 / 10 (avg.) | High (3/3 show awake more cost-effective long-term) | SD ± 15 – 20 % cost range (34.8 $k$ $vs$ 46.8 k; ICER – $82 k/ QALY) | Awake dominant economically and clinically (better KPS + QALY). | **B: Moderate** | Retrospective design downgrade 1 level; effect strong. |
| Colloid Cyst (Endoscopic vs Microsurgical) | Beaumont 2022 (USA single center) | High: same pathology, modern techniques | 7 / 10 | N/A (single study) | p = 0.007 and 0.0003 precise differences | Endoscopic cost ↓ by 52 %, LOS ↓ by 4 days, similar outcomes. | **C: Low-to-Moderate** | Single institution limits external validity. |
| Craniosynostosis (Endoscopic vs Open) | Garber 2017 (USA n = 300) | Moderate: pediatric only | 7 / 10 | High (consistency within subgroups) | SD acceptable ($21 k lowest) | Endoscopic dominant economically and clinically. | **B: Moderate** | Large sample improves confidence; generalizability limited to infants. |
| Vascular (MCA Aneurysm: open vs endovascular) | Lauzier 2023 (USA n = 136) | Moderate | 8 / 10 | Moderate: consistent within study only | Median cost 24 578 $v$ 39 737 | Endovascular dominant; no outcome loss reported | **C: Low-to-Moderate** | Retrospective single center downgrade. |
| Care Setting (Outpatient vs Inpatient Awa | Nassiri 2018 (Canada n = 50) | High: same indication (neoplasm) | 7 / 10 | High: parallel cost | Small sample – | Outpatient dominant economically with no | **C: Low-to-Moderat** | Pilot design; needs verification. |

| | | | | | | | |
|---|---|---|---|---|---|---|---|
| ke Craniotom y) | | | | saving signals | wide S D ( $5\ 242\ vs$ 7 225) | ↑ complications | **e** |

Economic evidence drawn from 11 clinical studies (19 145 patients) demonstrates a moderate-certainty association supporting the higher cost-effectiveness of craniotomy or minimally invasive cranial procedures compared with broader or more invasive alternatives. Confidence is strengthened by consistent directionality of cost and clinical outcomes across diverse settings (UK, USA, Finland, Cambodia), but downgraded for study-design heterogeneity and incomplete cost standardization across GBP, USD and EUR currencies.
